# Comprehensive care programmes in chronic obstructive pulmonary disease: a systematic review and meta-analysis of randomized controlled trials and real-world studies

**DOI:** 10.1101/2021.11.03.21265859

**Authors:** Daniel Yoo, Mengqi Gong, Lei Meng, Cheuk Wai Wong, Guangping Li, Michael Huen Sum Lam, Tong Liu, Gary Tse, Leonardo Roever, International Health Informatics Study (IHIS) Network

## Abstract

**Background:** Different comprehensive care programmes (CCPs) have been developed for patients with chronic obstructive pulmonary disorder (COPD), but data regarding their effectiveness have been controversial. PubMed and Embase were searched to 1^st^ June 2017 for articles that investigated the effects of the different types of CCPs on hospitalization or mortality rates in COPD.

**Results:** A total of 67 studies including 3472633 patients (mean age: 76.1±12.7 years old; 41% male) were analyzed. CCPs reduced all-cause hospitalizations (hazard ratio [HR]: 0.70, 95% confidence interval [CI]: 0.63-0.79; P<0.001; I^2^:96%) and mortality (HR: 0.69, 95% CI: 0.573-0.83; P<0.001; *I*^2^:75%). Subgroup analyses for different CCP types were performed. Hospitalizations were reduced by pharmacist-led medication reviews (HR: 0.54; 95% CI: 0.37-0.78; P=0.001; *I*^2^:49%), structured care programmes (HR: 0.76; 95% CI: 0.66-0.87; P<0.0001; *I*^2^:88%) and self-management programmes (HR: 0.79; 95% CI: 0.64-0.99; P<0.05; *I*^2^:78%), but not continuity of care programmes (HR: 0.70; 95% CI: 0.36-1.36; P=0.29; *I*^2^:100%), early support discharge or home care packages (HR: 0.97; 95% CI: 0.91-1.04; P=0.37; *I*^2^:0%) or telemonitoring (HR: 0.61; 95% CI: 0.32-1.18; P=0.14; *I*^2^:94%). Mortality was reduced by early support discharge or home care packages (HR: 0.49; 95% CI: 0.30-0.80; P<0.01; *I*^2^:72%), structured care programmes (HR: 0.69; 95% CI: 0.53-0.90; P<0.01; *I*^2^:61%) and telemonitoring (HR: 0.52; 95% CI: 0.31-0.89; P<0.05; *I*^2^:0%), but not self-management programmes (HR: 0.79; 95% CI: 0.64-0.99; P<0.05; *I*^2^:78%).

**Conclusions:** Comprehensive care programmes reduce hospitalization and mortality in COPD patients.

## Introduction

Chronic obstructive pulmonary disease (COPD) is a common, preventable and treatable disease that is characterized by persistent respiratory symptoms and airflow limitation that is due to airway and or parenchymal abnormalities usually caused by significant exposure to noxious particles or gases ^1^. It is a significant public health problem worldwide with an estimated prevalence between 11 and 26% in the BOLD Study Document “International variation in the prevalence of COPD”. It is also a significant cause of hospitalizations and mortality, placing a high burden on healthcare systems ^2-6^. These hospitalizations represent the single largest source of expenditure on patients with COPD ^7^. To tackle this, considerable efforts have focused on the development of comprehensive care programmes (CCPs) with the aim of reducing the number of hospitalizations and deaths. These include i) continuity of care programmes, ii) early support discharge with home care packages, iii) pharmacist-led medication reviews, iv) primary care programmes, and v) self-management programmes. However, not all studies have reported significant reductions in hospitalization or mortality following implementation of these programmes. Moreover, to date there has been no systematic analysis of the published literature to assess their benefits. Therefore, we performed a systematic review with meta-analysis of published studies on the effects of such programmes on hospitalization and mortality in COPD patients.

## Methods

### Search strategy, inclusion and exclusion criteria

This systematic review and meta-analysis was performed according to the Preferred Reporting Items for Systematic Reviews and Meta-Analyses statement ^8^. PubMed and Embase were searched up to 1^st^ June 2017, with no language restriction, for studies that investigated the hospitalization or mortality rates in COPD using the following terms: “((comprehensive care) AND COPD) AND (hospitalization or readmission or mortality or death)”. The following inclusion criteria were applied: i) the design was a case-control, prospective or retrospective cohort study or RCT in humans, ii) hazard ratios with 95% confidence intervals (CI) for all-cause hospitalization or mortality rates were reported or could be calculated from the published data.

Quality assessment of cohort studies included in our meta-analysis was performed using the Newcastle–Ottawa Quality Assessment Scale (NOS) (**Supplementary Table 1** for case-control studies, **Supplementary Table 2** for cohort studies) ^9^, and of RCTs using the Cochrane Risk of Bias Tool (**Supplementary Figures 1 and 2**). The NOS evaluated the categories of study participant selection, comparability of the results, and quality of the outcomes. The following characteristics were assessed: a) representativeness of the exposed cohort; b) selection of the non-exposed cohort; c) ascertainment of exposure; d) demonstration that outcome of interest was not present at the start of study; e) comparability of cohorts on the basis of the design or analysis; f) assessment of outcomes; g) follow-up period sufficiently long for outcomes to occur; and h) adequacy of follow-up of cohorts. This scale varied from zero to nine stars, which indicated that studies were graded as poor quality if they met <5 criteria, fair if they met 5 to 7 criteria, and good if they met >8 criteria.

### Data extraction and statistics

Data from the different studies were entered in pre-specified spreadsheet in Microsoft Excel. All potentially relevant entries were retrieved as complete manuscripts and assessed for compliance with the inclusion criteria. In this meta-analysis, the extracted data elements consisted of: i) publication details: last name of first author, publication year and locations; ii) study design (case-control study, cohort study or RCT); iii) follow-up duration; iv) endpoints; v) the quality score; and vi) the characteristics of the population including sample size, gender, age and number of subjects. Meta-analyses of observational studies are challenging due to differences in study designs and inherent biases. Two reviewers (GT and CC) independently reviewed each included study and disagreements were resolved by adjudication with input from a third reviewer (TL).

The endpoints for this meta-analysis were hospitalization and mortality rates. Multivariate adjusted hazard ratios (HRs) or relative risks (RRs) with 95% CI were extracted and analyzed for each study. When values from multivariate analysis were not available, those from univariate analysis were used. When the latter were not provided, raw data were used to calculate unadjusted risk estimates where possible. The pooled adjusted risk estimates from each study as the OR values with 95% CI were presented.

Heterogeneity between studies was determined using Cochran’s Q, the weighted sum of squared differences between individual study effects and the pooled effect across studies, and the *I*^*2*^ statistic from the standard chi-square test, which is the percentage of the variability in effect estimates resulting from heterogeneity. *I*^*2*^ > 50% was considered to reflect significant statistical heterogeneity. A fixed effects model was used if *I*^*2*^ < 50%. The random-effects model using the inverse variance heterogeneity method was selected if *I*^*2*^ > 50%. To locate the origin of the heterogeneity, sensitivity analysis by the leave-one-out method was performed. Subgroup analyses based on the type of care programmes were performed. Where a study reported effective estimates at successive time points, the longest time point was used. Funnel plots, Begg and Mazumdar rank correlation test and Egger’s test were used to assess for possible publication bias.

## Results

A flow diagram detailing the search strategy and study selection process is shown in **Figure 1**. A total of 855 and 57 studies were retrieved from PubMed and Embase, respectively. Of these, 68 were included in the final meta-analysis. The baseline characteristics of these studies are listed in **Table 1**. Five were case-control studies, 36 were cohort studies and 27 were randomized controlled trials. The mean follow-up duration was 47 months. In this study, 3472633 patients were analyzed (mean age: 76.1 ± 12.7 years old; 41% male).

**Figure 1.**
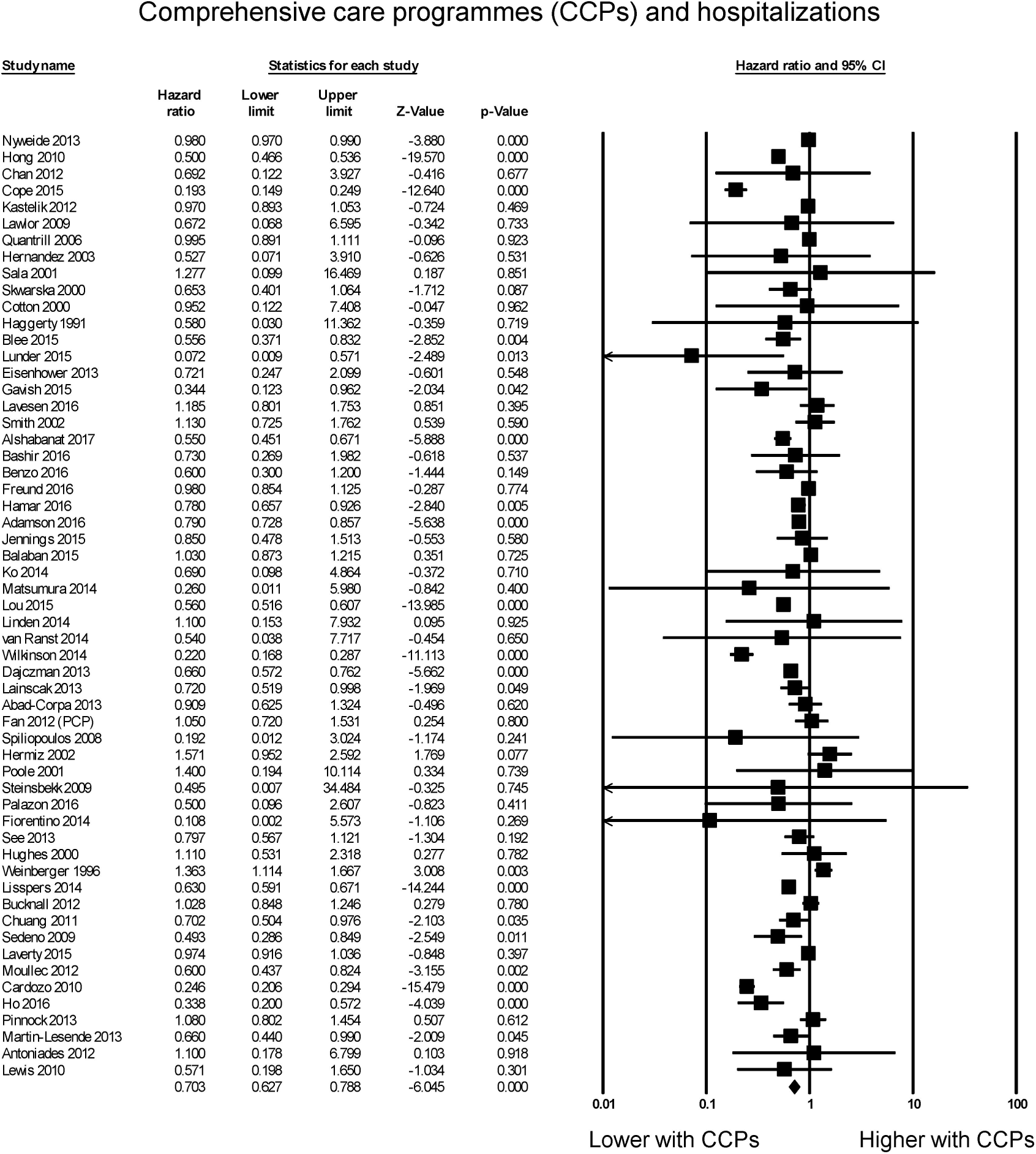
A flow diagram detailing the search strategy and study selection process for this systematic review and meta-analysis on the effects of comprehensive care programmes on hospitalization and mortality rates in chronic obstructive pulmonary disease (COPD).

### CCPs and all-cause hospitalizations

The effectiveness of CCPs compared to usual care was evaluated in 67 studies. Of these, 20 reported significant reductions, 36 demonstrated no significant difference and one study reported significant increase in all-cause hospitalizations. The overall meta-analysis showed that CCPs reduced all-cause hospitalizations by 30% (hazard ratio: 0.70, 95% confidence interval: 0.63-0.79; P < 0.001; **Figure 2**). *I*^2^ took a value of 96%, indicating the presence of substantial heterogeneity. Sensitivity analysis by leaving out one study at a time did not significantly alter the pooled HR (**Supplementary Figure 3**). Funnel plot plotting standard errors against the logarithms of the hazard ratio are shown in **Supplementary Figure 4**. Begg and Mazumdar rank correlation demonstrated no significant publication bias (Kendal’s Tau value = 0.09, P > 0.05) but Egger’s test demonstrated significant asymmetry (intercept -2.1, t-value 3.3; *P* < 0.05).

**Figure 2.**
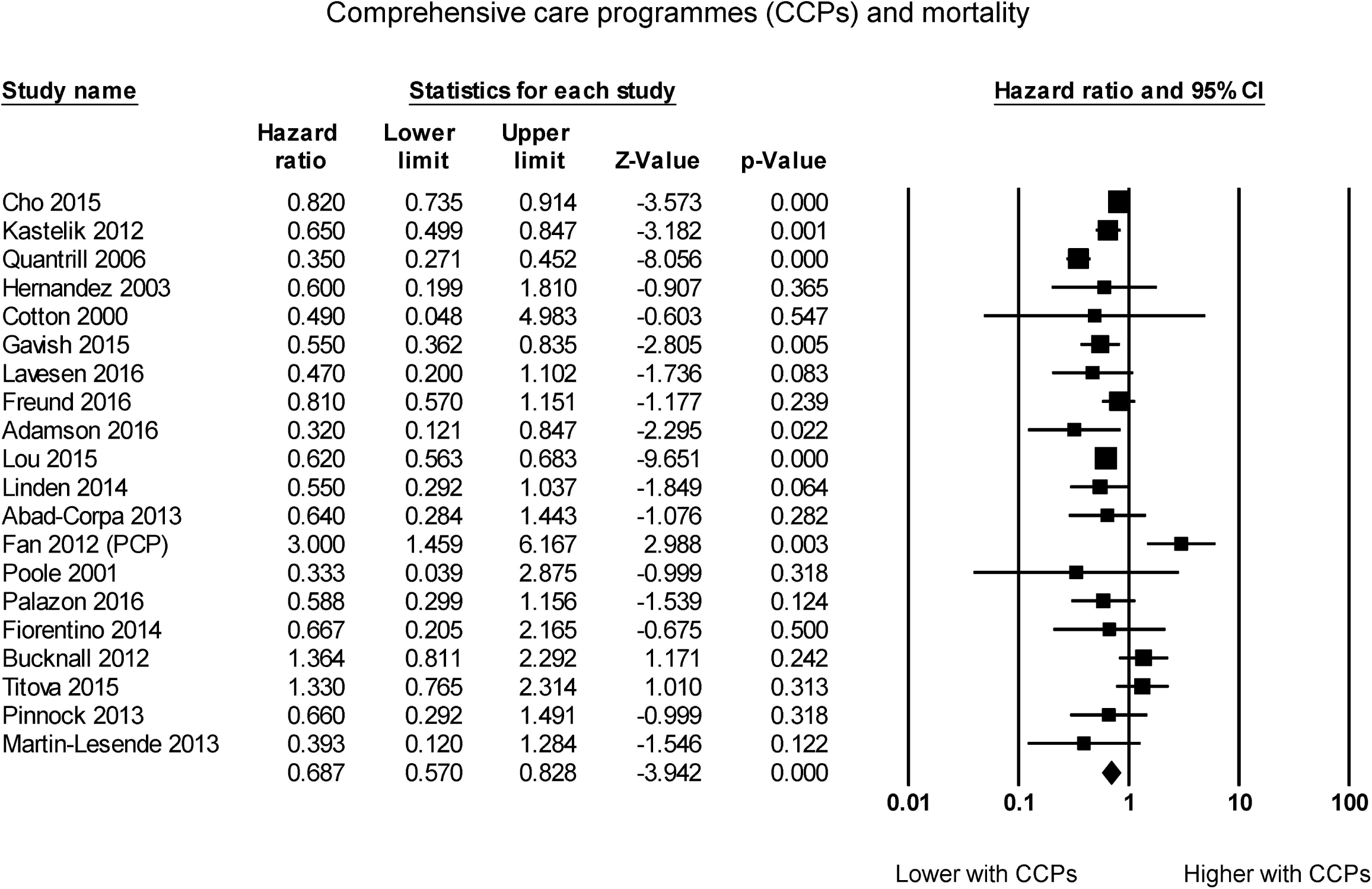
Pooled hazard ratios for studies examining the effects of comprehensive care programmes on hospitalization rates in COPD.

Subgroup analysis for the effects of different CCPs on hospitalization rates were performed. Continuity of care programmes (HR: 0.70; 95% CI: 0.36-1.36; P = 0.29; *I*^2^ = 100%; **Supplementary Figure 5**) or early support discharge or home care package (HR: 0.97; 95% CI: 0.91-1.04; P = 0.37; *I*^2^ = 0%; **Supplementary Figure 6**) or telemonitoring (HR: 0.61; 95% CI: 0.32-1.18; P = 0.14; *I*^2^ = 94%; **Supplementary Figure 7**) did not significantly reduce hospitalizations. By contrast, pharmacist-led medication review (HR: 0.54; 95% CI: 0.37-0.78; P = 0.001; *I*^2^ = 49%; **Supplementary Figure 8**), structured care programmes (HR: 0.76; 95% CI: 0.66-0.87; P < 0.0001; *I*^2^ = 88%; **Supplementary Figure 9**) and self-management programmes (HR: 0.79; 95% CI: 0.64-0.99; P < 0.05; *I*^2^ = 78%; **Supplementary Figure 10**) all significantly reduced hospitalizations.

### CCPs and all-cause mortality

The effectiveness of CCPs compared to usual care on all-cause mortality was also examined. CCPs significantly reduced mortality rates by 31% (HR: 0.69, 95% CI: 0.573-0.83; P < 0.001; **Figure 3**). *I*^2^ took a value of 75%, indicating the presence of substantial heterogeneity. Sensitivity analysis by leaving out one study at a time did not significantly alter the pooled HR (**Supplementary Figure 11**). A funnel plot of standard errors against the logarithms of the hazard ratio are shown in **Supplementary Figure 12**. Begg and Mazumdar rank correlation demonstrated no significant publication bias (Kendal’s Tau value = -0.03, P > 0.05). Egger’s test demonstrated no significant asymmetry (intercept -0.06, t-value 0.1; *P* > 0.05).

**Figure 3.**
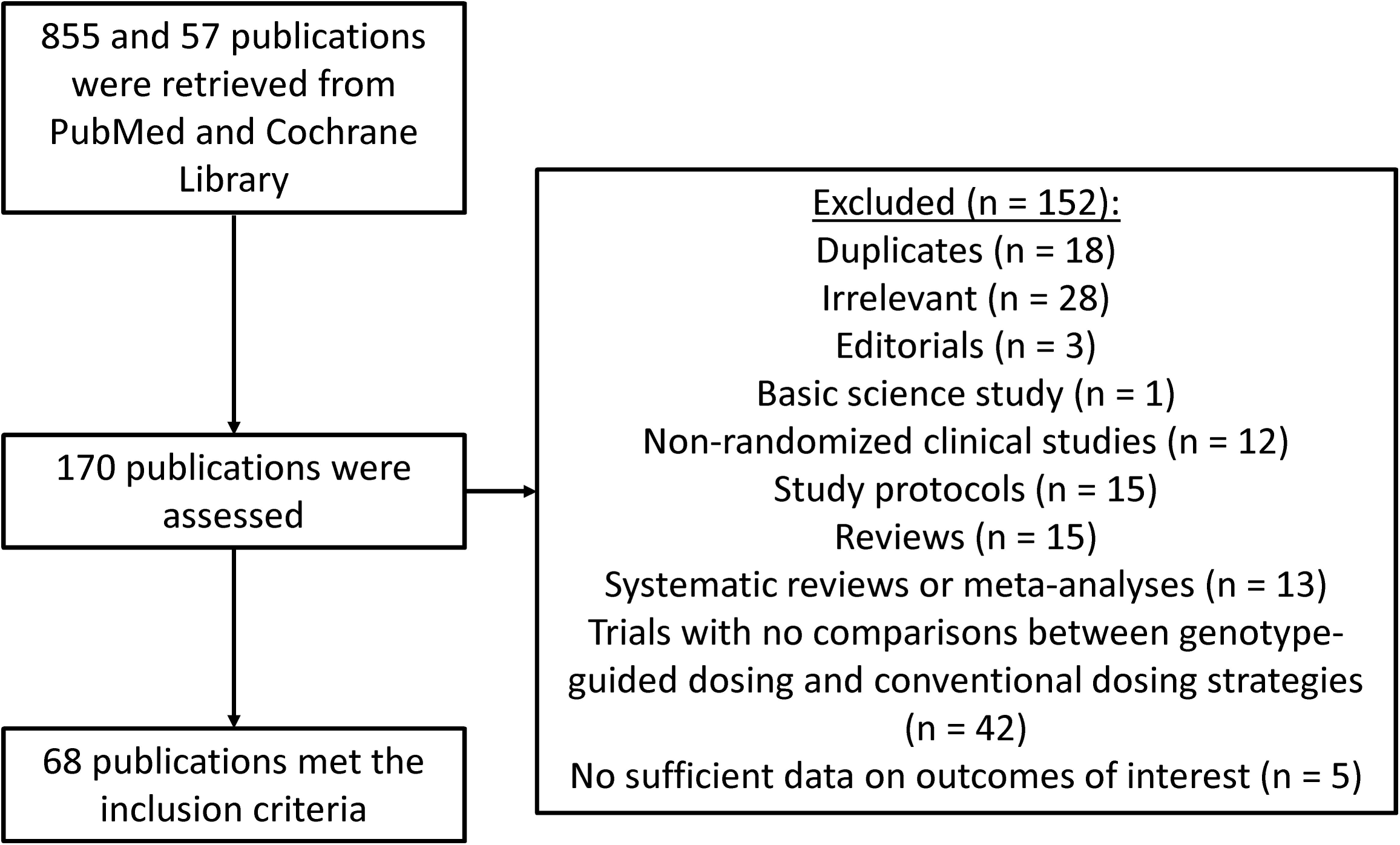
Pooled hazard ratios for studies examining the effects of comprehensive care programmes on mortality rates in COPD.

Subgroup analyses were performed for the effects of different types of CCPs on mortality. Early support discharge or home care package (HR: 0.49; 95% CI: 0.30-0.80; P < 0.01; *I*^2^ = 72%; **Supplementary Figure 13**), structured care programmes (HR: 0.69; 95% CI: 0.53-0.90; P < 0.01; *I*^2^ = 61%; **Supplementary Figure 14**) and telemonitoring (HR: 0.52; 95% CI: 0.31-0.89; P < 0.05; *I*^2^ = 0%%; **Supplementary Figure 15**) significantly reduced mortality, whereas self-management programmes did not (HR: 1.35; 95% CI: 0.92-1.97; P = 0.12; *I*^2^ = 0%; Supplementary Figure 16).

## Discussion

We conducted a systematic review and meta-analysis of RCTs and real-world studies on the effects of CCPs on hospitalization and mortality rates in COPD. The main findings are that CCPs reduced (1) hospitalization rates by 30% and (2) mortality by 31%.

COPD is a complex disease that requires input from different members of the multidisciplinary team. Different formal CCPs have been designed to provide integrated care to the patients, but their effectiveness is controversial. A Cochrane review conducted in 2013 demonstrated improvement in disease-specific quality of life, the number of hospital admissions, inpatient length-of-stay but not mortality ^10^. Moreover, well-organized care is important for good clinical outcomes but this needs not necessarily involve integrated programs, as shown by a randomized controlled trial that compared integrated disease management programmes with usual care in a healthcare system that has good support networks ^11^. In our systematic review, we identified different types of CCPs, and broadly divided them into the following of continuity of care programmes, early support discharge or home care package, pharmacist-led medication review, primary care programmes, self-management programmes, and telemonitoring.

### Continuity of care programmes

Fragmented visit patterns may be related to preventable hospitalizations and conversely continuity of care programmes may be able to reduce hospitalizations. Different tools are available to quantify continuity of care (COC). Of these, the COC Index features both total number of providers and the total number of ambulatory care visits ^12^. A recent retrospective cohort study examined the relationship between COC index and avoidable hospitalizations in COPD patients ^13^. Regarding the effects of COC on mortality, one study reported a significant reduction in mortality ^14^, whereas another study reported no relationship between COC index and mortality ^15^. From our meta-analysis, there was insufficient evidence that continuity of care programmes reduced hospitalizations and it was not possible to perform meta-analysis of two included studies for hazard ratios on mortality due to lack of reporting of outcome data in one study ^15^. Nevertheless, a recently published systematic review and meta-analysis of randomized controlled trials specifically on COC programmes demonstrated a significant reduction in COPD-related, but not all-cause, hospital readmission rates ^16^.

### Early support discharge or home care package

Early support discharge programmes, as the name suggests, involves early discharge of the patient with COPD followed by a home care package involving education, smoking cessation advice, chest physiotherapy, home exercise with contact details provided to the patients. The aim is to reduce the number of hospitalizations and inpatient length-of-stay without compromising patient safety. A systematic review and meta-analysis of randomized controlled trials published in 2003 reported that hospital readmission and mortality rates for patients receiving early support discharge programmes were comparable to those receiving inpatient care ^17^. Our study provides further evidence that early support discharge or home care package was safe, and has the benefit of significantly reducing mortality in COPD patients. However, the hospital readmission rates were not significantly different from those for standard care, which is in keeping with the finding of the previously published meta-analysis on hospital-at-home care ^17^. Moreover, one of the largest single study on patients offered supported discharge programmes reported significant reduction in mortality compared to those not receiving such care. This may be due to the fact that only the patients who are less sick were enrolled into the early discharge programmes whereas the more severe cases were directed towards inpatient care. Together, our findings support the notion that early support discharge programmes are safe and cost-effective, through decreasing the inpatient length-of-stay and from patient-centered point of view since patients and their carers prefer domiciliary care over hospital care ^18^. However, this must be done cautiously, and risk stratification is important to identify those who will likely to benefit from such programmes.

### Pharmacist-led medication review

Medication discrepancies have been identified as a risk factor for unplanned hospital readmission especially in elderly patients ^19^ and these are significantly reduced by pharmacist input ^20^. The involvement of a pharmacist in the discharge medication reconciliation can aid identification of discrepancies in medications and in the context of COPD, has been shown to reduce hospital readmissions. In COPD, adherence to prescribed medications including inhalers is important and a pharmacist-educator model has been shown to improve compliance ^21^. Our meta-analysis shows that pharmacist-led medication review was effective in reducing hospitalizations.

### Self-management programmes

Self-management programmes aims to help patients develop coping mechanisms, maintain active lifestyle, promote drug compliance and encourage early identification of symptoms to prevent exacerbations. A Cochrane systematic review and meta-analysis of randomized and non-randomized controlled trials demonstrated improvements in health-related quality of life and dyspnea symptoms and a reduction in respiratory-related hospital admissions, but not all-cause hospital readmissions or mortality ^22^. Interestingly, our meta-analysis showed that self-management programmes significantly reduced all-cause hospitalizations. These findings suggest that self-management education can help patients recognize exacerbation symptoms, facilitate early treatment and prevent hospitalization. Moreover, our study showed that mortality was not significantly increased or decreased, thereby confirming the safety of these self-management programmes in COPD patients.

### Telemonitoring

Telemonitoring refers to the use of telecommunication technology to transmit data on vital signs and symptoms of patients, and medications to a centrally located operator who will relay the information to a clinician, who can modify treatment options with the aim of preventing deterioration ^23^. Since the aim of this study was not to examine the effectiveness of telemonitoring, and the search terms did not include strings on telemonitoring, only a small number of studies on the effects of telemonitoring on outcomes of COPD patients were identified. In our meta-analysis, telemonitoring did not significantly reduce hospitalizations but was effective in reducing mortality. By contrast, a systematic review of randomized and non-randomized controlled trials published in 2014 on the effectiveness of telemonitoring reported significant reductions in COPD exacerbations and readmissions ^24^. A similar study published in 2014 also reported significant reductions in hospitalizations whilst reporting a non-statistical increase in mortality ^25^. Contrastingly, our meta-analysis demonstrated a reduction in mortality although this may be due to a small number of studies included.

### Structured care programmes

In our systematic review, the programmes that did not fit into the above categories were pooled together as structured care programmes. The latter can take on many forms, such as a pre-defined care plan involving different members of the multidisciplinary team of a doctor, nurse, physiotherapist, occupational therapist, social workers and dieticians ^26^. This may focus on different domains such as initial assessment, evaluation of functional and activities of daily living, patient education, smoking cessation advice, nutritional strategies. As an example, case management programmes have been tested for their efficacy in reducing healthcare utilization and mortality in COPD. Some included studies also used a case management approach. Case management is defined by the American Case Management Association (ACMA) in 1986 as a “comprehensive and cooperative process, which includes assessment, planning, implementation, cooperation, quality control, and service evaluation to determine the level of patient satisfaction with health care provided”. This typically involves a case manager who are allocated patients and see through their care process post-discharge from the hospital ^27^. Our meta-analysis has demonstrated reductions in both hospitalizations and mortality with the implantation of structural care programmes for COPD patients.

## Limitations

There are several limitations of this study that must be recognized. Firstly, HRs of RCTs and cohort studies, which are different study designs, were pooled together. This may be justified as a recent Cochrane review showed that there were no significant difference in the effective estimates between observational studies and RCTs, suggesting that factors other than study design are responsible for differences in outcomes ^28^. Secondly, there was a high level of heterogeneity observed when all of the studies were pooled together. This probably reflected the heterogeneous nature of the different CCPs. However, we were only partly successful in reducing the heterogeneity on our subgroup analysis. These findings suggest that other factors, such as follow-up duration, or patient characteristics, may have produced the heterogeneity.

## Conclusion

This meta-analysis demonstrates that comprehensive care programmes are effective for reducing hospitalizations and mortality in COPD patients.

## Data Availability

All data produced in the present work are contained in the manuscript

## Author contributions

GT: conception of study, data extraction, analysis, interpretation, manuscript drafting and critical revision, supervision of study

AY, CWW, MG, LM: data extraction, analysis, interpretation, manuscript drafting

GL, MHSL, TL: data interpretation, critical revision of manuscript, supervision of study

LR: conception of study, data interpretation, critical revision of manuscript, supervision of study

GT agrees to be the guarantor of the paper, taking responsibility for the integrity of the work as a whole, from its inception to published article.

