## Supplementary material for "Comprehensive care programmes in chronic obstructive pulmonary disease: a systematic review and meta-analysis of randomized controlled trials and real-world studies": Table 1

**Table 1** Characteristics of the 12 studies on hemodynamic monitoring included in this meta-analysis.

| First author / Year | Study design | Type of comprehensive care programme | Sample size (n) | Age | SD | % Male | Follow-up (months) |
| --- | --- | --- | --- | --- | --- | --- | --- |
| Cho 2015 | Cohort | Continuity of care | 3090 | 69.0 | 10.1 | 76 | 84 |
| Lin 2015 | Cohort | Continuity of care | 3015 | - | - | 65 | 12 |
| Nyweide 2013 | Cohort | Continuity of care | 3276635 | 76.4 | 10.8 | 41 | 48 |
| Hong 2010 | Cohort | Continuity of care | 131512 | 72.1 | 5.1 | 46 | 48 |
| Chan 2012 | Cohort | DNIV | 132 | 71.2 | 16.3 | 84 | 12 |
| Cope 2015 | Cohort | ESD/HCP | 9 | - | - | - | 12 |
| Kastelik 2012 | Cohort | ESD/HCP | 9716 | 72.0 | 10.0 | - | 3 |
| Lawlor 2009 | Cohort | ESD/HCP | 246 | 67.4 | 12.7 | - | 12 |
| Quantrill 2006 | Cohort | ESD/HCP | 3511 | 72.0 | 10.4 | 52 | 3 |
| Hernandez 2003 | RCT | ESD/HCP | 222 | 70.8 | 9.7 | 97 | 2 |
| Sala 2001 | Cohort | ESD/HCP | 205 | 68.0 | 14.9 | - | 0.5 |
| Skwarska 2000 | RCT | ESD/HCP | 184 | 70.6 | - | 47 | 2 |
| Cotton 2000 | RCT | ESD/HCP | 81 | 66.8 | 2.0 | 43 | 2 |
| Haggerty 1991 | Cohort | ESD/HCP | 20 | 66.0 | 7.8 | 53 | 37 |
| Blee 2015 | RCT | Medication | 620 | 62.0 | 11.0 | 54 | 2 |
| Lunder 2015 | Prospective | Medication | 90 | 66.1 | 17.5 | 39 | 1 |
| Eisenhower 2013 | Prospective | Medication | 29 | 74.2 | 9.3 | 44 | 1 |
| Gavish 2015 | Retrospective Cohort | PCP | 195 | 66.0 | 10.0 | 83 | 3 |
| Lavesen 2016 | RCT | PCP | 213 | 70.2 | 14.2 | 39 | 3 |
| Smith 2002 | Cohort | PCP | 377 | - | - | - | 12 |
| Alshabanat 2017 | Cohort | PCP | 1564 | 73.9 | 17.2 | 54 | 24 |
| Ko 2017 | RCT | PCP | 180 | 74.7 | 8.2 | 96 | 12 |
| Bashir 2016 | Case-control | PCP | 461 | 70.9 | 13.4 | 67 | 1 |
| Benzo 2016 | RCT | PCP | 215 | 68.0 | 13.4 | 45 | 12 |
| Freund 2016 | RCT | PCP | 2076 | 72.0 | 13.6 | 48 | 24 |
| Hamar 2016 | Cohort | PCP | 1505 | 71.9 | 13.2 | 42 | 6 |
| Adamson 2016 | Case-control | PCP | 462 | 70.6 | 18.6 | 62 | 3 |
| Jennings 2015 | RCT | PCP | 172 | 64.7 | 15.1 | 45 | 3 |
| Balaban 2015 | RCT | PCP | 1510 | 64.7 | 22.8 | 42 | 1 |
| Ko 2014 | RCT | PCP | 185 | 76.9 | 7.4 | 90 | 12 |
| Matsumura 2014 | Cohort | PCP | 11 | 75.9 | 4.8 | 100 | 24 |
| Lou 2015 | RCT | PCP | 8217 | 61.5 | 18.9 | 48 | 48 |
| Linden 2014 | RCT | PCP | 512 | 67.7 | 11.8 | 42 | 3 |
| van Ranst 2014 | Cohort | PCP | 34 | 72.0 | 9.0 | 49 | 12 |
| Wilkinson 2014 | Cohort | PCP | 202 | - | - | - | 12 |
| Dajczman 2013 | Cohort | PCP | 253 | 71.8 | 9.7 | 50 | 13 |
| Lainscak 2013 | RCT | PCP | 143 | 71.0 | 9.0 | 72 | 6 |
| Abad-Corpa 2013 | RCT | PCP | 426 | 72.8 | 11.4 | 91 | 6 |
| Fan 2012 (PCP) | RCT | PCP | 363 | 66.0 | 11.7 | 97 | 8 |
| Spiliopoulos 2008 | Cohort | PCP | 50 | 74.9 | - | 46 | 54 |
| Lu 2007 | RCT | PCP | 50 | 73.6 | 18.1 | 76 | - |
| Hermiz 2002 | RCT | PCP | 133 | 66.9 | - | 47 | 3 |
| Poole 2001 | Case-control | PCP | 32 | 72.7 | - | 59 | 12 |
| Steinsbekk 2009 | Cohort | PCP | 30 | 62.8 | 8.5 | 44 | 36 |
| Ries 1995 | RCT | PCP | 119 | 62.6 | 10.2 | 73 | 72 |
| Palazon 2016 | Cohort | PCP | 100 | 73.0 | 9.2 | 68 | - |
| Fiorentino 2014 | Case-control | PCP | 72 | - | - | - | 12 |
| See 2013 | Cohort | PCP | 123 | - | - | - | 1 |
| Hughes 2000 | RCT | Primary care | 1966 | 70.4 | 14.6 | 96 | 12 |
| Weinberger 1996 | RCT | Primary care | 1396 | 62.8 | 15.6 | 99 | 6 |
| Lisspers 2014 | Cohort | Primary care | 16404 | 68.0 | 16.2 | 47 | 12 |
| Bucknall 2012 | RCT | SMP | 464 | 69.1 | 9.3 | 37 | 12 |
| Chuang 2011 | Case-control | SMP | 282 | - | - | - | 12 |
| Sedeno 2009 | RCT | SMP | 166 | 69.3 | 9.8 | 56 | 12 |
| Bourbeau 2003 | RCT | SMP | 191 | 69.5 | 9.8 | 55 | 12 |
| Laverty 2015 | Cohort | SMP + PCP | - | - | - | - | 1 |
| Titova 2015 | Prospective cohort | SMP + PCP | 199 | 73.4 | 9.3 | 43 | 24 |
| Moullec 2012 | Retrospective cohort | SMP + PCP | 189 | 72.1 | 14.7 | 50 | 12 |
| Fan 2012 (SMP + PCP) | RCT | SMP + PCP | 426 | 66.0 | 11.7 | 97 | 8 |
| Cardozo 2010 | Cohort | Telemonitoring | 851 | - | - | 32 | 2 |
| Ho 2016 | RCT | Telemonitoring | 106 | 80.2 | 8.8 | 76 | 6 |
| Pinnock 2013 | RCT | Telemonitoring | 256 | 68.9 | 12.2 | 45 | 12 |
| Martin-Lesende 2013 | RCT | Telemonitoring | 58 | 81.0 | 7.5 | 59 | 12 |
| Antoniades 2012 | RCT | Telemonitoring | 44 | 69.0 | 12.0 | 45 | 12 |
| Lewis 2010 | RCT | Telemonitoring | 40 | 68.5 | 13.5 | 50 | 6 |

Abbreviations: RCT: randomized controlled trial; DNIV: domiciliary non-invasive ventilation; ESD/HCP: Early support discharge/home care package; PCP: Pharmacist-led care programme; SMP: self-management programme
