## Supplementary Appendix for "Comprehensive care programmes in chronic obstructive pulmonary disease: a systematic review and meta-analysis of randomized controlled trials and real-world studies"

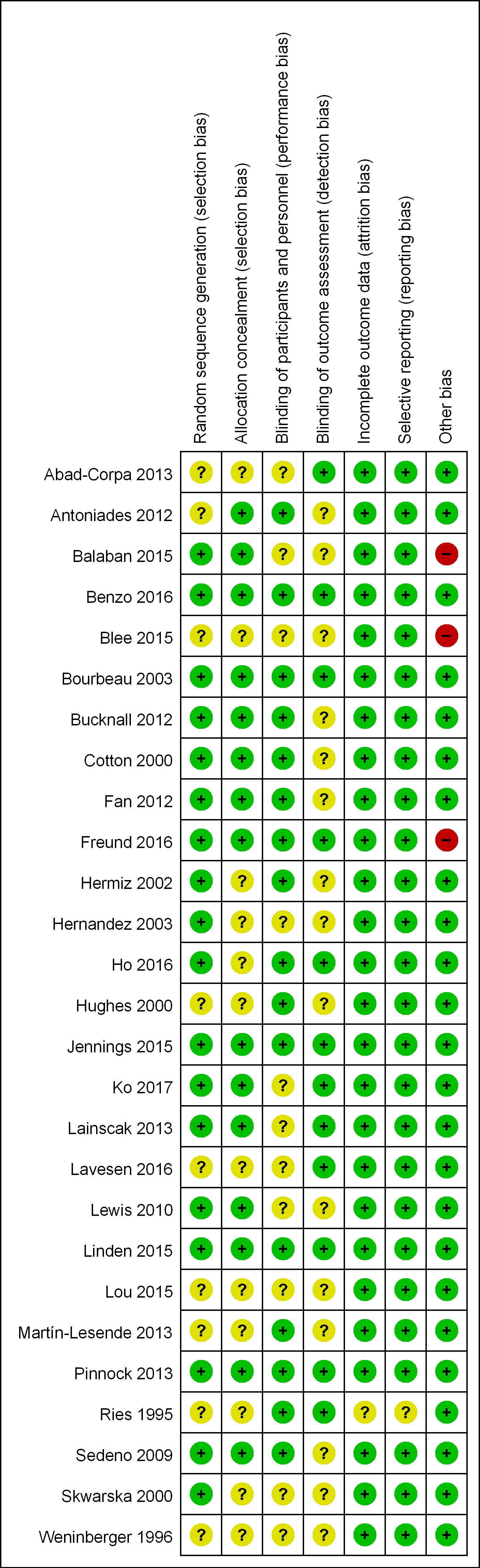

Supplementary Figure 1. Cochrane Risk of Bias Tool analysis for individual randomized controlled trials.

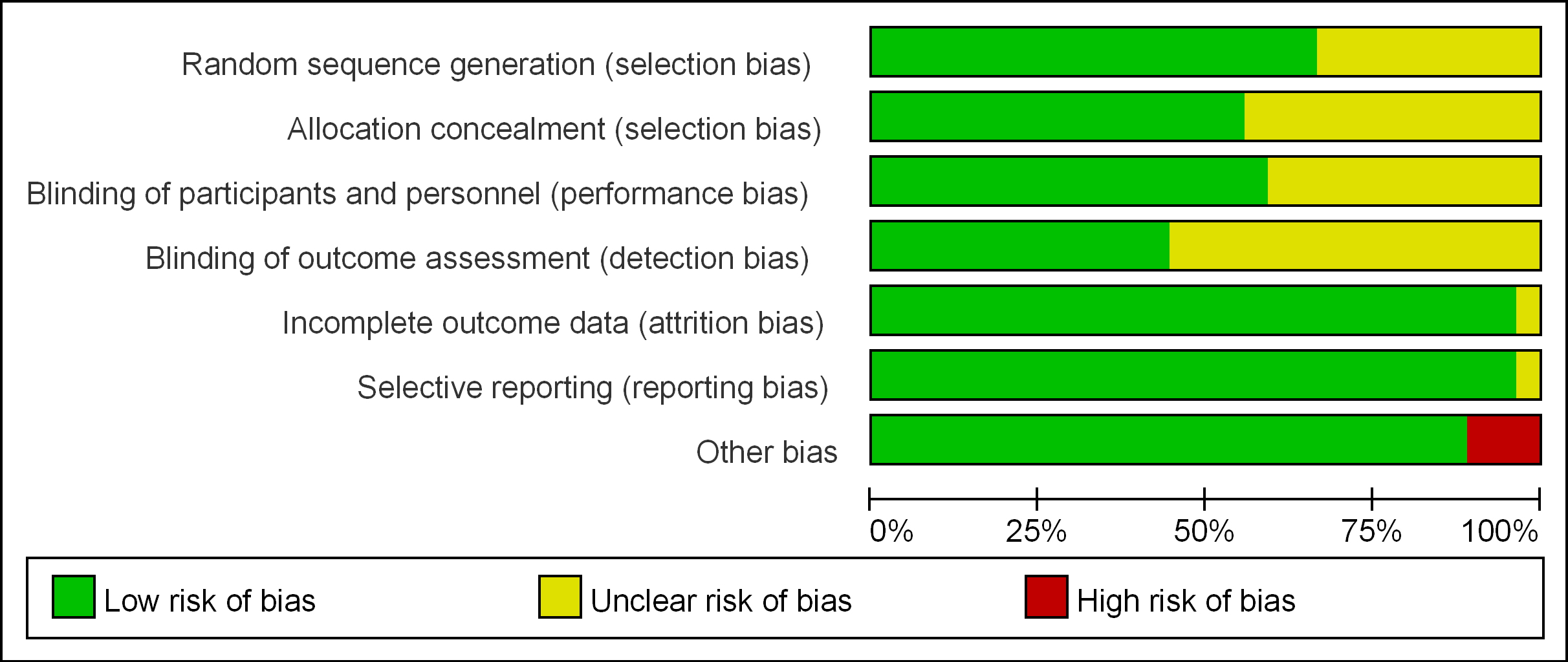

Supplementary Figure 2. Cochrane Risk of Bias Tool summary for all randomized controlled trials.

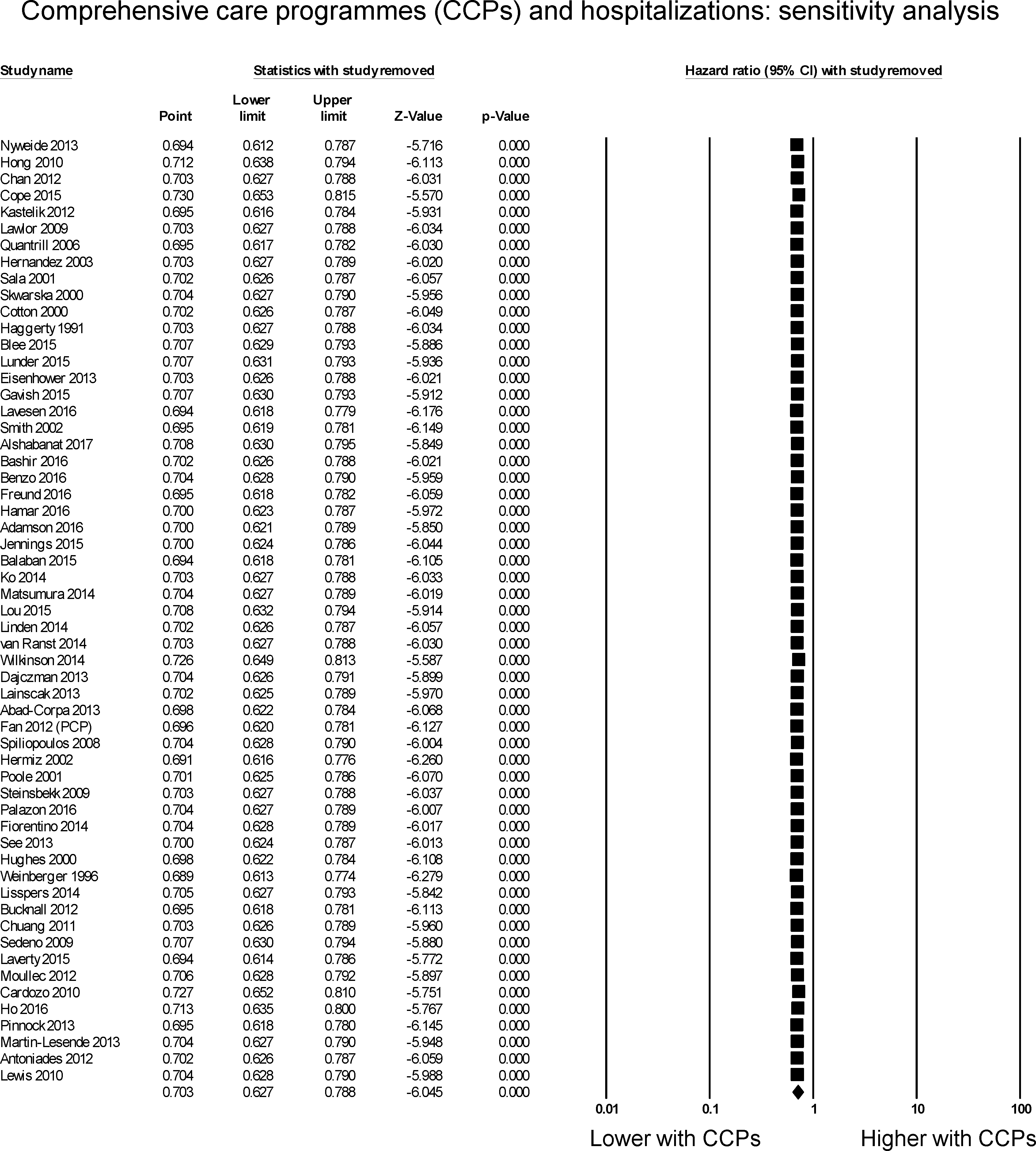

Supplementary Figure 3. Sensitivity analysis by leaving out one study at a time for hazard ratios studies examining the effects of all comprehensive care programmes on hospitalization rates in COPD.

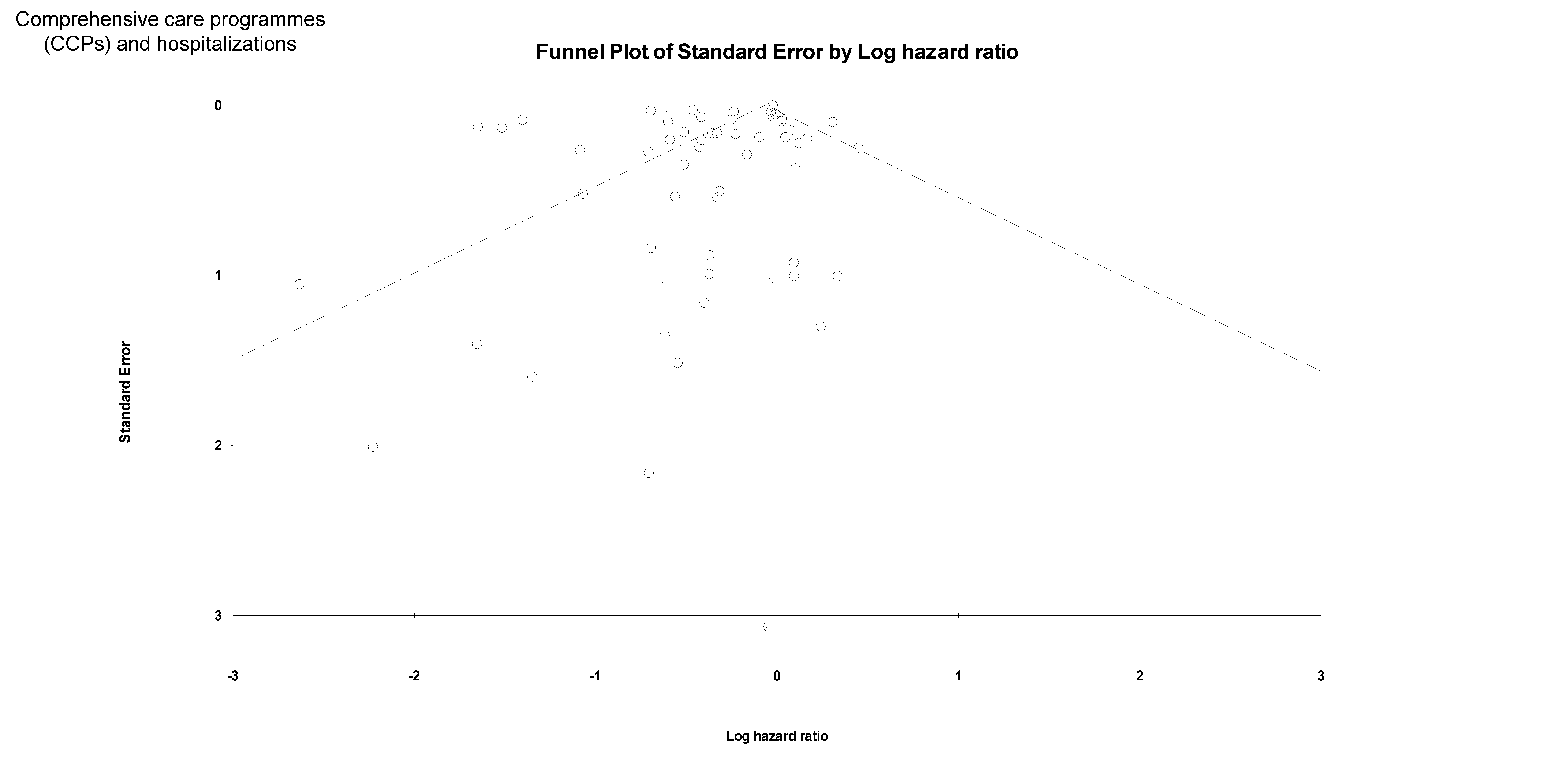

Supplementary Figure 4. Funnel plots of standard error against the logarithm of hazard ratios for comprehensive care programmes and hospitalization rates in COPD.

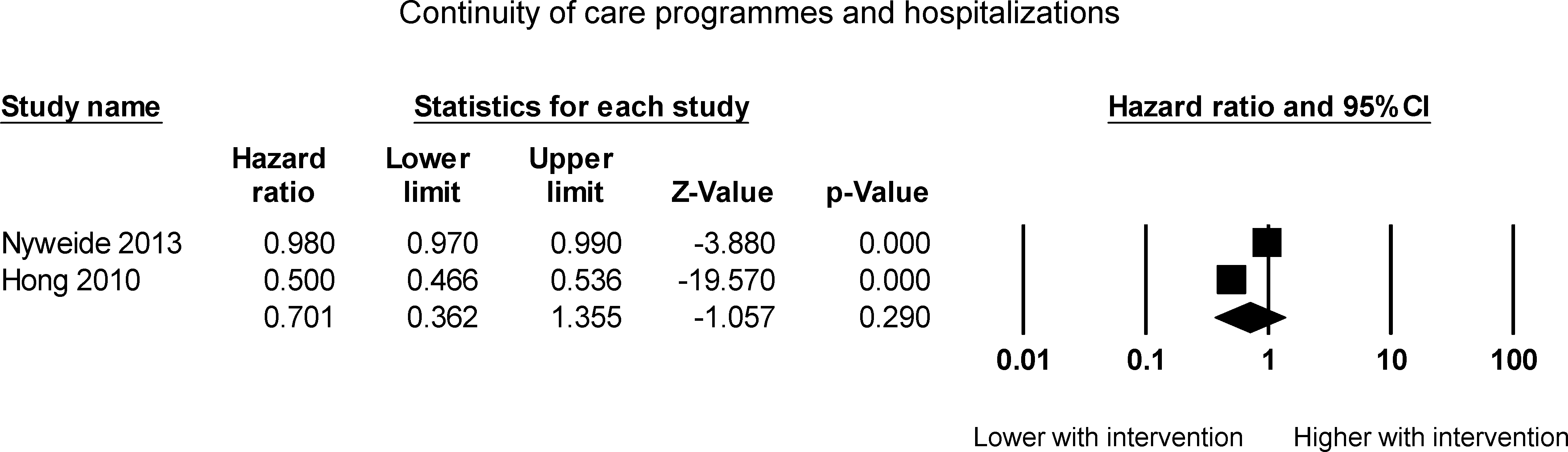

Supplementary Figure 5. Pooled hazard ratios for studies examining the effects of continuity of care programmes on hospitalization rates in COPD.

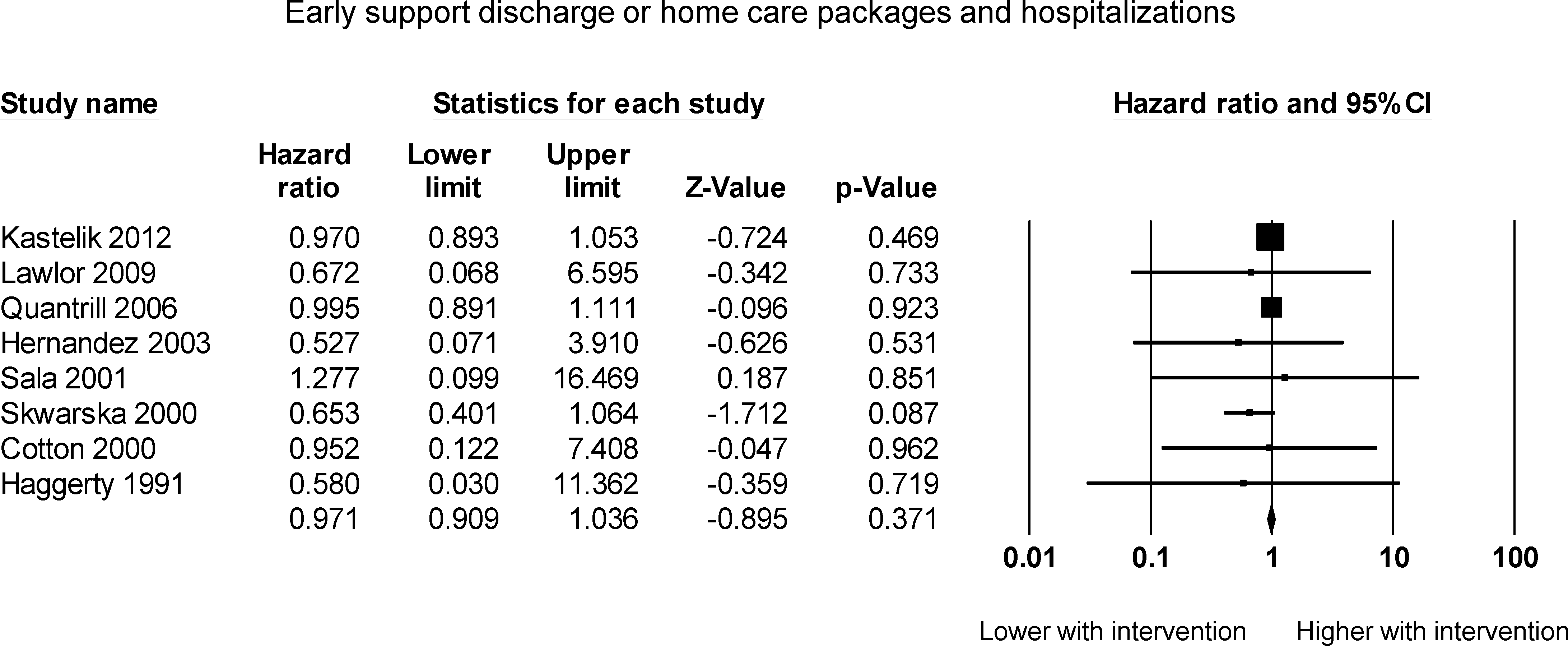

Supplementary Figure 6. Pooled hazard ratios for studies examining the effects of early support discharge programmes or home care packages on hospitalization rates in COPD.

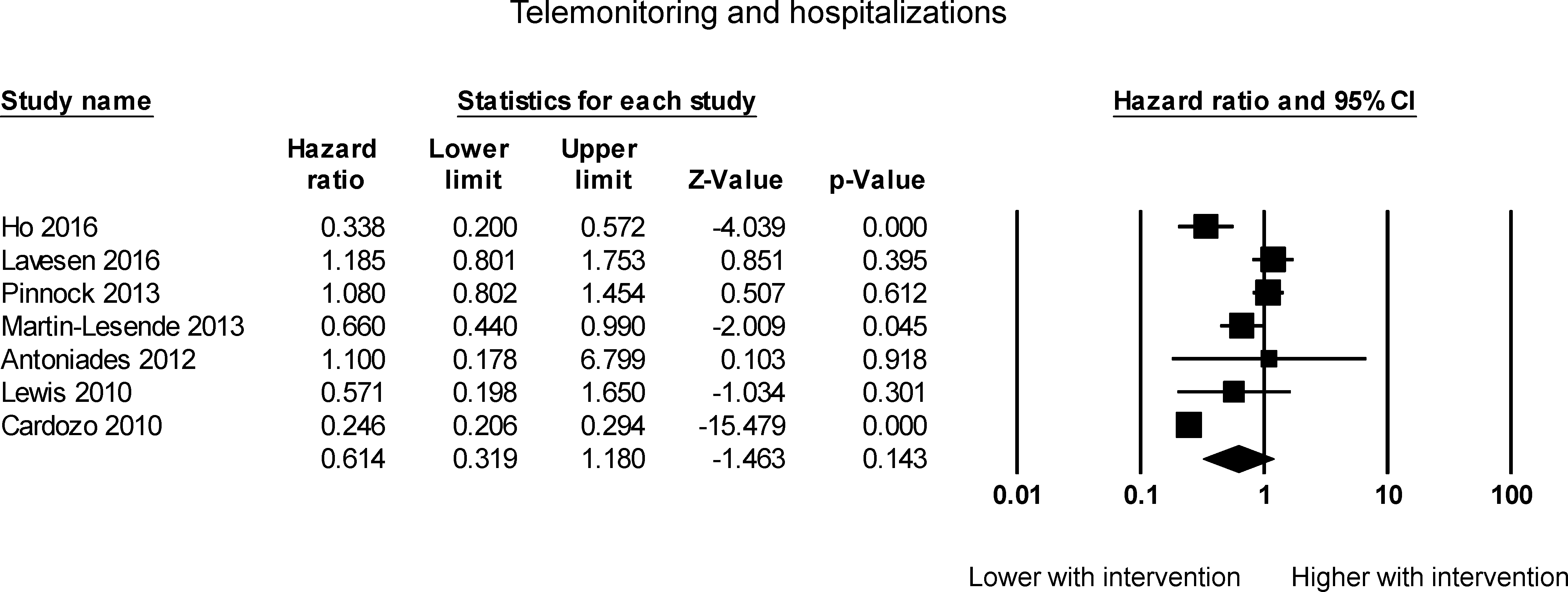

Supplementary Figure 7. Pooled hazard ratios for studies examining the effects of telemonitoring on hospitalization rates in COPD.

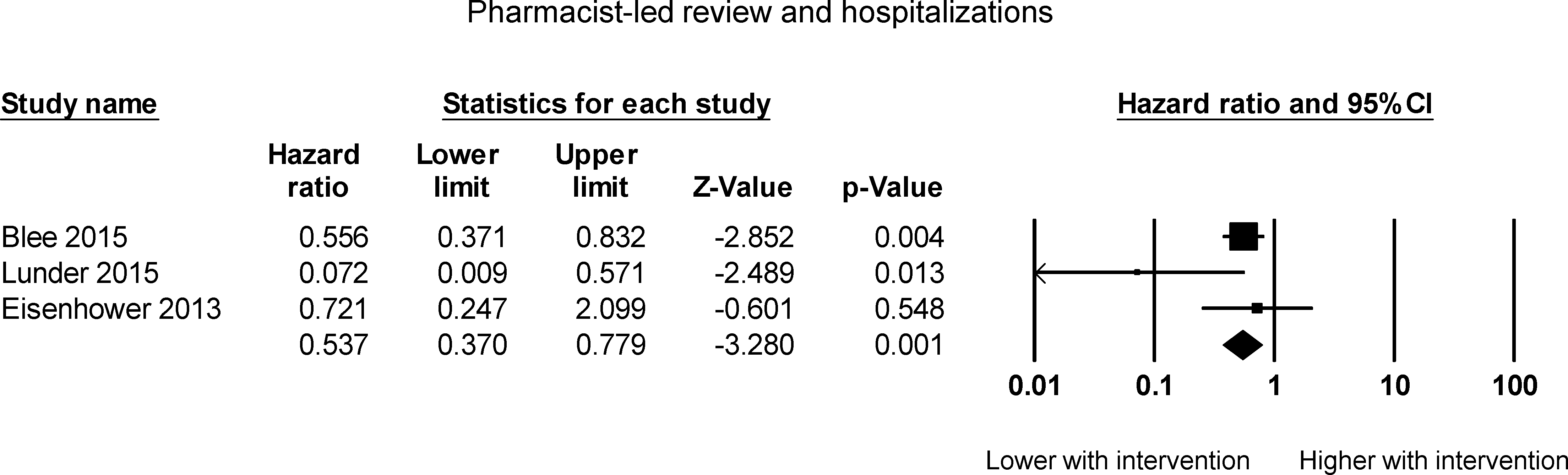

Supplementary Figure 8. Pooled hazard ratios for studies examining the effects of pharmacist-led reviews on hospitalization rates in COPD.

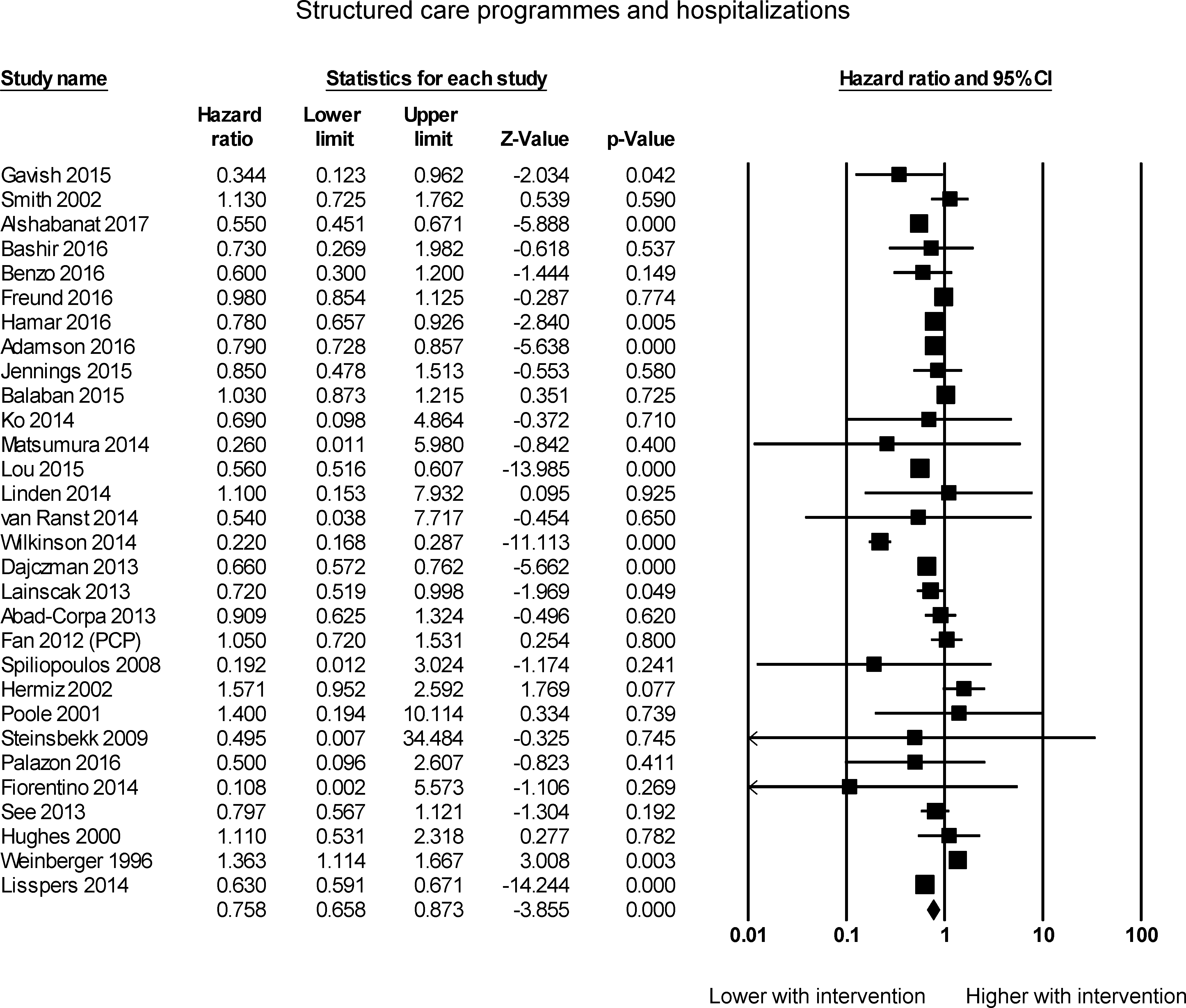

Supplementary Figure 9. Pooled hazard ratios for studies examining the effects of structured care programmes on hospitalization rates in COPD.

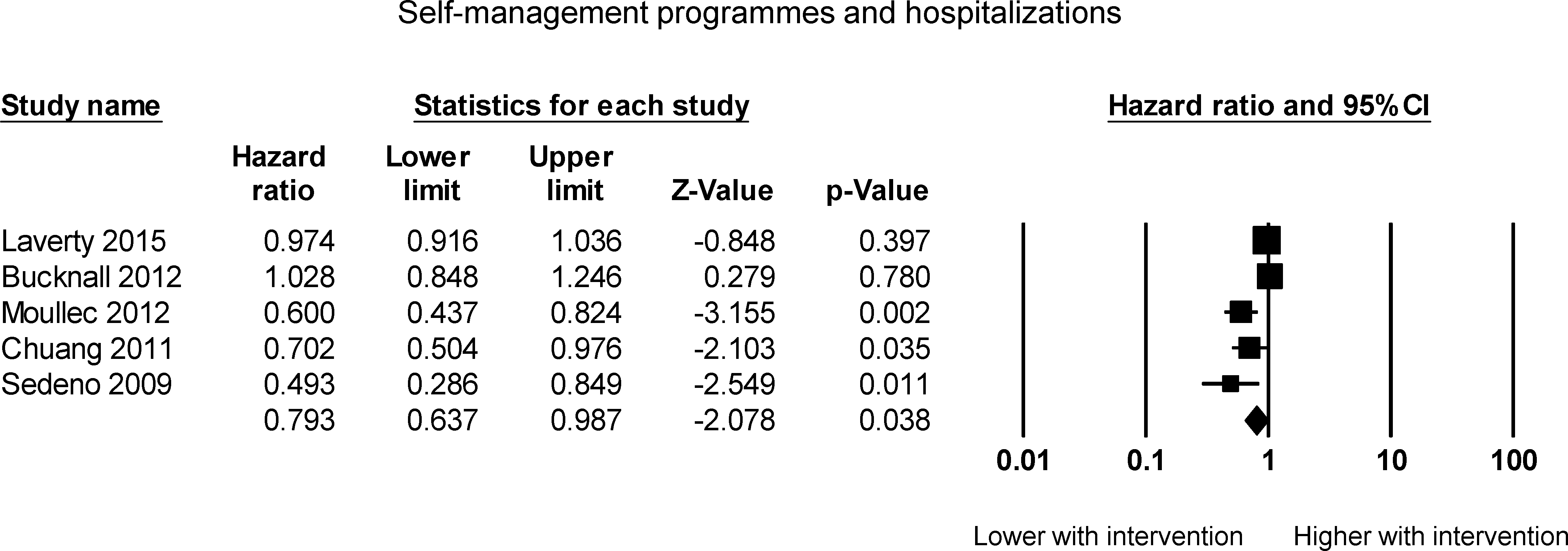

Supplementary Figure 10. Pooled hazard ratios for studies examining the effects of self-management programmes on hospitalization rates in COPD.

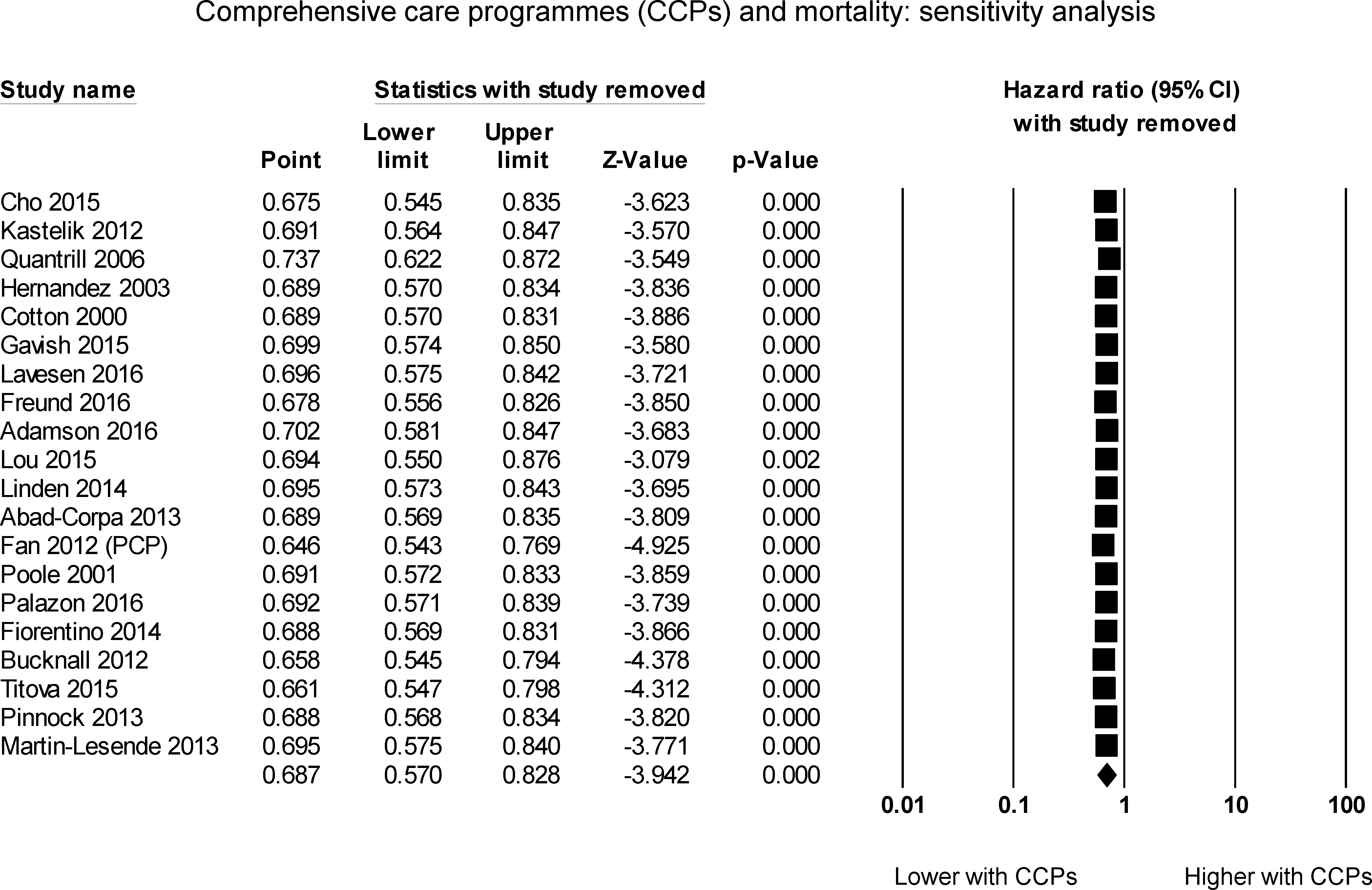

Supplementary Figure 11. Sensitivity analysis by leaving out one study at a time for hazard ratios studies examining the effects of all comprehensive care programmes on mortality rates in COPD.

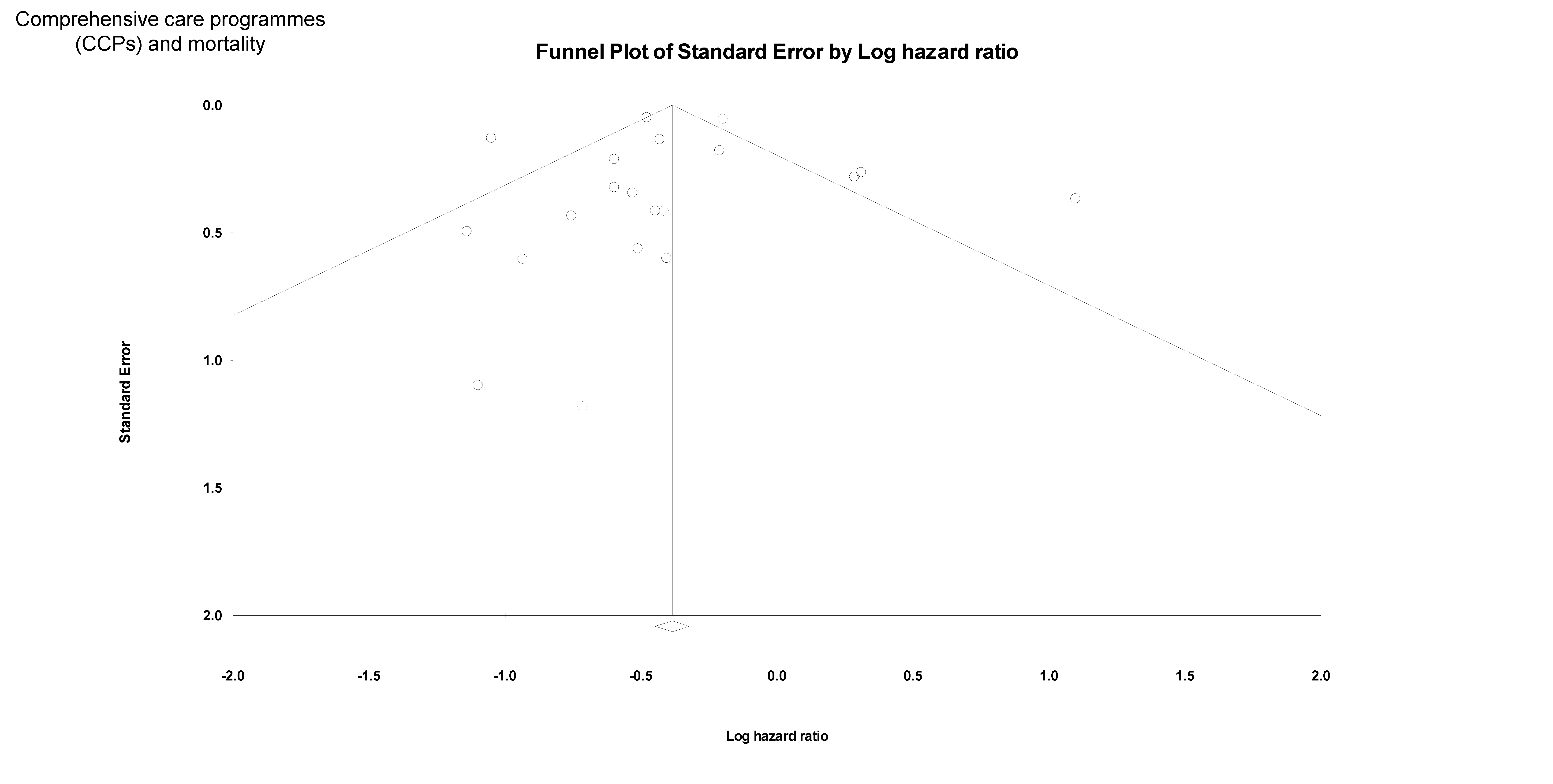

Supplementary Figure 12. Funnel plots of standard error against the logarithm of hazard ratios for comprehensive care programmes and mortality rates in COPD.

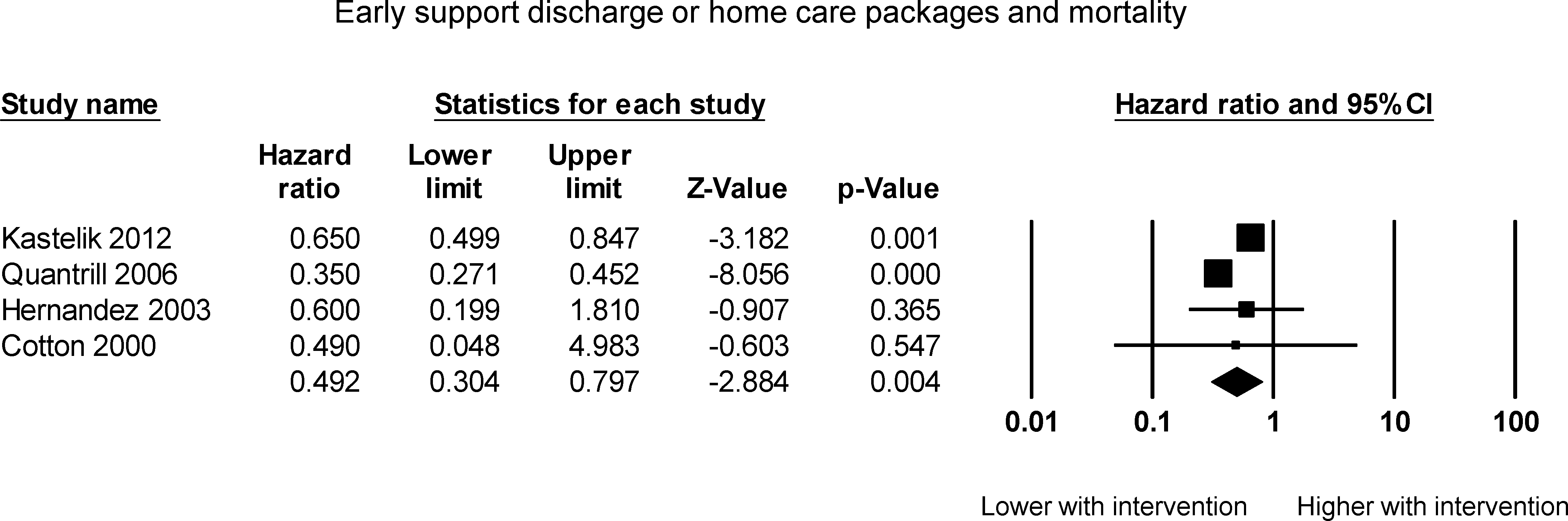

Supplementary Figure 13. Pooled hazard ratios for studies examining the effects of early support discharge and home care packages on mortality rates in COPD.

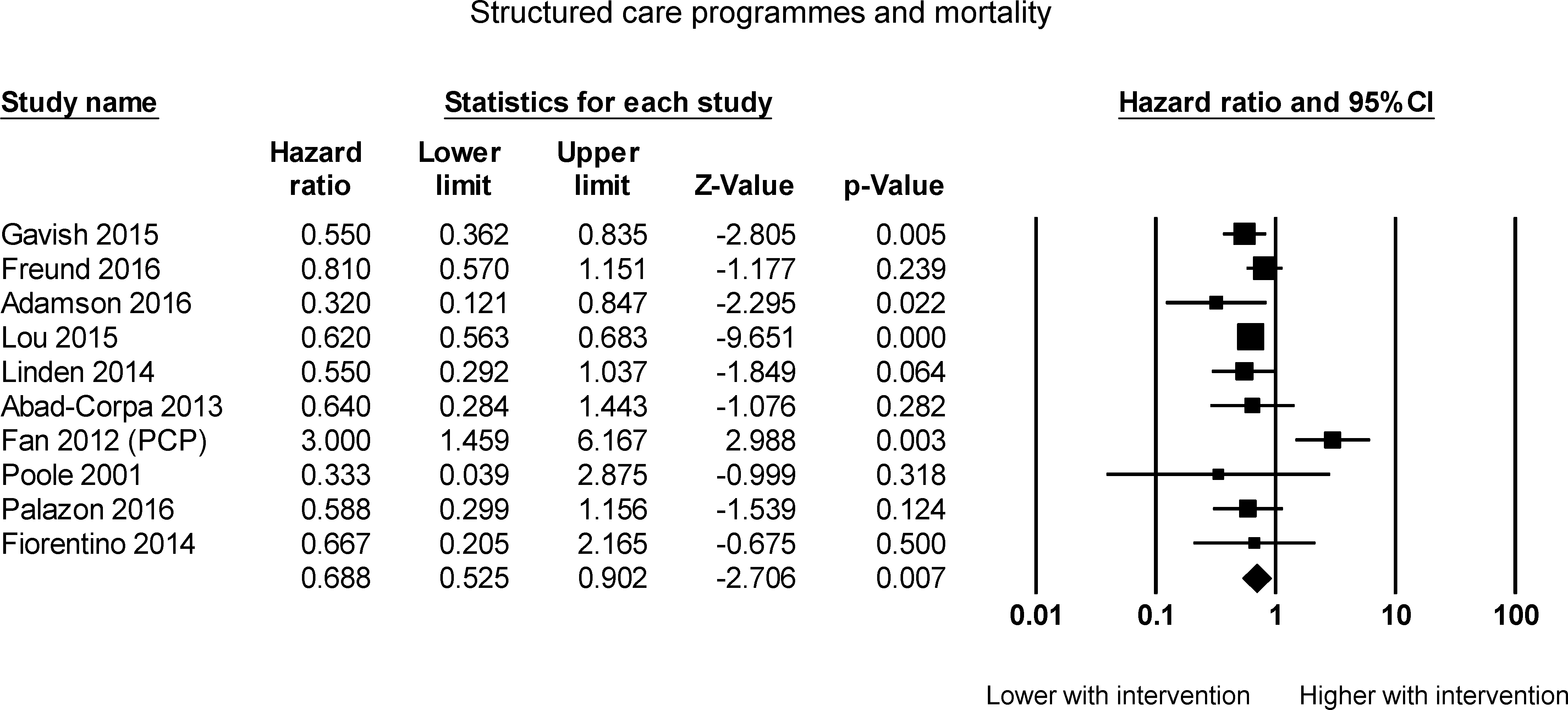

Supplementary Figure 14. Pooled hazard ratios for studies examining the effects of structured care programmes on mortality rates in COPD.

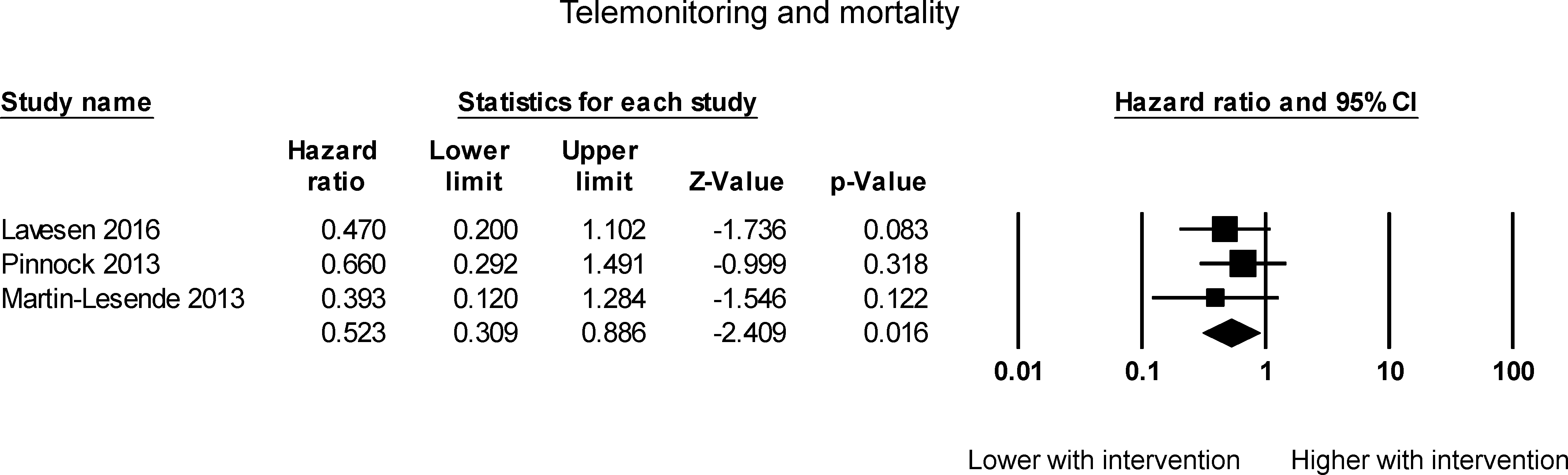

Supplementary Figure 15. Pooled hazard ratios for studies examining the effects of telemonitoring on mortality rates in COPD.

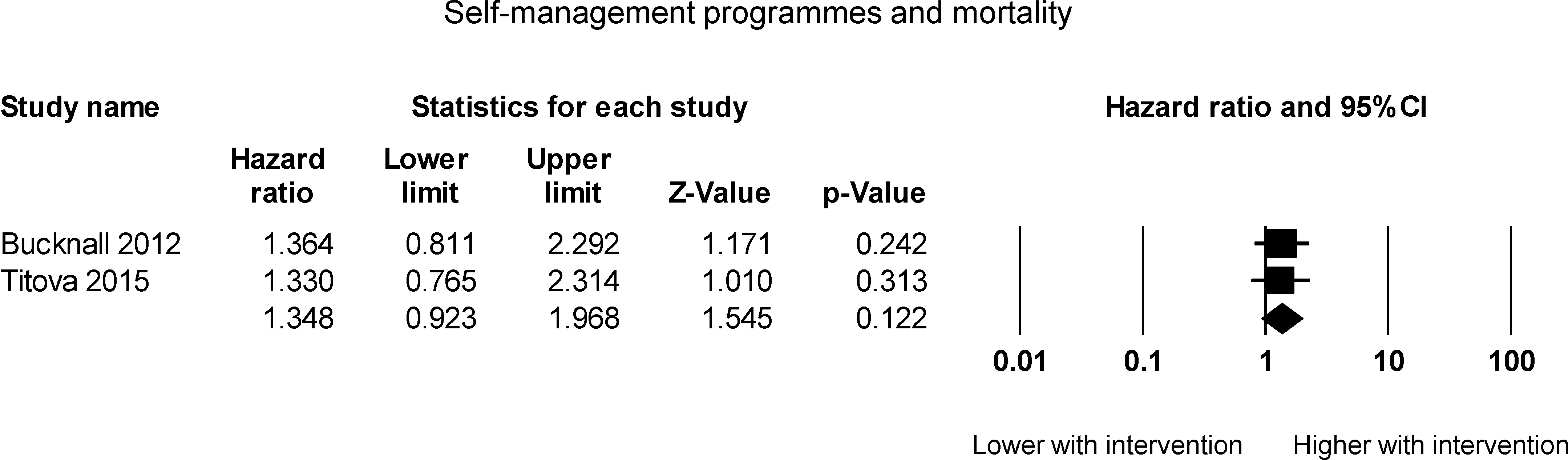

Supplementary Figure 16. Pooled hazard ratios for studies examining the effects of self-management programmes on mortality rates in COPD.
